# Introducing and Testing Maternal Vulnerability Segmentation Tool (MVST) for Essential Service Utilization: Initial Evidence from Implementation Research in Oromia, Ethiopia

**DOI:** 10.1101/2024.12.28.24319732

**Authors:** Yihunie Lakew, Habtamu Tamene, Bee-Ah Kang, Nandita Kapadia-Kundu, S. H Kuka, Rajiv Rimal

## Abstract

In Ethiopia, utilization of maternal health services, including antenatal care and health facility delivery remain low. This is particularly acute among women living with individual, social, and environmental vulnerabilities. This study aimed to develop and test a maternal vulnerability segmentation tool to enable community workers to identify pregnant women least likely to access maternal care services. Guided by Project Pathways’ Vulnerability Framework, predictors of maternal health service utilization were identified, and a 20-item questionnaire was developed from identified predictors. We employed exploratory and confirmatory factor analysis to create a latent variable called maternal vulnerability. Vulnerability items with acceptable internal reliability were tested to confirm association with antenatal care and institutional delivery through mediation regression analysis. A factor with eigenvalue greater than one was identified, which were further reduced and refined to 10, four for urban-rural residents, and six for a rural sample. This resulted in a root mean square error of approximation (RMSEA) values of between 0.01 to 0.08, comparative fit index (CFI) > 0.90, and Tucker Lewis index (TLI) value > 0.90. Women with higher vulnerability score were less likely to access antenatal care visits and more likely to deliver at home (*p*<0.01). Our data-driven work leveraging this tool provide guidance to effective programs in identifying underprivileged pregnant women to enable community workers to narrow maternal health service gaps in Ethiopia.

**What is already known on this topic:**

- Nearly half of pregnant women are not using essential maternal services such as antenatal care and health facility delivery, placing their delivery and own health at risk.
- Pregnant women are least likely to use those services because of individual, social, economic, and environmental factors

**What this study adds:**

- This study provides new maternal vulnerability segmentation tools to identify women living with vulnerabilities for targeted interventions. It explains the extent to which intersecting vulnerability scores affect essential maternal service utilization.

**How this study might affect research, practice or policy:**

- This study enables community health workers to classify pregnant women according to their degree of vulnerability, thus paving the way for developing well-targeted interventions
- It also calls for further research aimed at generating more validated vulnerability segmentation tools that effectively and efficiently address maternal vulnerabilities across different contexts in low-income and middle-income countries.

## Introduction

Maternal and child mortality has been a priority for the global agenda ^1^, targeting under the 3^rd^ Sustainable Development Goal to reduce maternal mortality ratio to less than 70 per 100,000 live births, neonatal mortality to lower than 12 deaths per 1,000 live births, and under-5 mortality to at least as low as 25 deaths per 1,000 live births by 2030 ^2^. Around 38% aggregate mortality reduction has been documented between 2000-2017, but the progress is uneven across geographic and population segments ^3^ due to various context-specific factors ^4–6^.

Ethiopia has shown progress in many maternal and child health indicators over the past few decades. In 2000, the maternal mortality ratio (MMR) was 871 deaths per 100,000 live births and declined to 412 in 2016 ^7^. Despite this substantial progress, the country faces persistent health challenges. MMR accounts for 3.6% of global maternal mortality ^2^. Twenty-five percent of female deaths were due to pregnancy-related causes ^7^, and the neonatal mortality rate also increased by four points per 1,000 livebirths between 2016 and 2019 ^8^.

Women’s use of health facilities for childbirth (a predictor of MMR) greatly varies by region and sociodemographic characteristics. With the national average being approximately 48%, the percentage of institutional delivery ranges from 23.3% in Somali Region to 94.8% in Addis Ababa, the capital city ^8^. The percentage of health facility delivery in Oromia region is 41%, lower than the national average ^8^. Also, 79% of women in the lowest wealth quintile delivered a baby at home; the corresponding figure for women in the highest wealth quintile was 14%. This implies appropriate action is required to address such disproportionate uptake of and access to maternal health services. To achieve faster and more equitable improvements, the government of Ethiopia has implemented a three-tiered health system with an intensive health extension program (a community-based approach) to improve health literacy and access to primary healthcare ^9^.

Maternal and child health (MCH) services, including antenatal care and health facility delivery, are critical in reducing mortality ^10^. Accessing such services before and during labor can significantly reduce the risk of maternal and neonatal deaths attributable to prematurity, intrapartum, or postpartum complications ^11–13^. For example, timely access to maternal child health services has been shown to reduce maternal deaths by between 16-33% ^14^ and neonatal mortality by 29% ^15^.

In Ethiopia, there are significant disparities in access to essential services, implying that strategies must be tailored to the specific geographic, social, and health contexts in each region and segments of population. One such strategy is to develop a vulnerability segmentation tool, which identifies factors that put women at risk and then tailor interventions accordingly. Vulnerability segmentation is becoming an important concept in such efforts ^4,16^. Despite its importance, less attention has been paid to how vulnerability is defined, conceptualized, and operationalized in the realm of maternal health service coverage ^16^. There is also a lack of consensus on what constitutes vulnerability and the underlying multidimensional factors that comprise it ^16,17^.

Substantial literature exists on vulnerabilities in different geographic, economic, environmental, and social contexts, which manifest as disparities in maternal and newborn health outcomes. In extant literature, vulnerability has two schools of thought: the behavioral perspective, and the structuralist perspective ^18^, though there are also other perspectives, including external and internal sides of vulnerability, with external referring to risk, and internal referring to individual capacities for coping ^19^. In the economics literature, vulnerability is conceptualized in terms of poverty dynamics, food security, or sustainable livelihoods ^19^. In recent years, resilience has also emerged as a similar concept, with it referring to how individuals cope with the stressors and outcomes they are subjected to as a result of their vulnerabilities ^20^. Furthermore, several vulnerability indexes are available, including those measuring economic, socio-environmental, drought, and heat-related vulnerabilities ^21–23^.

Though of great utility, these indexes are primarily intended for national and sub-national use, and their applicability to community household or individual levels are limited. For example, in the United States, there is a maternal vulnerability index (MVI) available at county and state levels ^24^. It was developed by Surgo Ventures and has 43 indicators across six themes: (i) reproductive health care, (ii) physical health, (iii) mental health and substance abuse, (iv) general health care, (v) socio-economic determinants and (vi) physical environment. Although the MVI is useful for identifying geographics with high or low maternal risk, it cannot identify households and individuals using those multidimensional vulnerability drivers ^24^. MVI’s application includes research to show the index’s association with adverse health outcomes, which generates evidence of the association between vulnerability and poor maternal outcomes. Several other maternal vulnerability scales have also been developed, but they are not designed for household and individual level ^22–25^. Several scoping reviews define vulnerability in the context of maternal and child health with factors broadly categorized into three themes: socioeconomic, biological, and environmental ^26,27^. In this study, vulnerability is defined in terms of risk exposure and adaptive behaviors, referring to underserved pregnant women affected by diverse social, environmental, and behavioral factors. Although the majority of women and child population is vulnerable, evidence revealed that there is a lack of thorough understanding of the socio-ecological factors to leverage the reduction of their vulnerability ^28^. Vulnerability assessment tools should address both individual and systemic risk exposures in addition to a “system’s ways and means of coping” ^29^.

Segmenting pregnant women by degree of vulnerability enhances the scope of providing equitable maternal health services and in a cost-effective manner. By tailoring strategies specific to the segmented audience, we envision interventions to use precious resources in the most effective manner. The rationale of this study, then, is to identify pregnant women who are at higher risk of home delivery and use an intervention tailored to their circumstances.

In the literature, there is a lack of guidance on how to identify specific individuals with maternal vulnerability at the community level so that frontline health workers can address their specific needs. Therefore, the objectives of the study are three-fold: (1) to develop maternal vulnerability segmentation tools for essential service utilization from implementation research field practice data, (2) to develop maternal vulnerability indexes, (3) to test if relationships exist between degree of vulnerability and essential maternal service utilization.

## Methods

Data for this study, collected at baseline before the intervention, comes from a community-based quasi-experiment run in Oromia, Ethiopia in early 2024. The study was designed as a pre- and post-intervention among a panel sample of pregnant women. Within this region, two woredas were selected as the treatment arm and two others as control arm. The treatment arm received the intervention, whereas the control did not (it was conceptualized as usual care). Data collection modalities and frequencies were the same in both arms: we collected data at baseline, before the start of the intervention and then again at end-line, after the intervention. Details are indicated elsewhere ^30^.

At baseline, to establish a sampling frame and to reach pregnant women with vulnerabilities, a total of 1,614 households were visited through house-to-house visit using a simple pregnancy screening tool to get suspected pregnancies and resulted in 1189 pregnant women with greater or equal to five months of pregnancy. All of those pregnant women were listed eligible for vulnerability segmentation and interviewed in a face-to-face interview using 20-item questionnaire, automated in a digital data collection tool. After listing eligible pregnant women, we randomly sampled three women per enumeration area to achieve our target sample size. If an enumeration area had five or fewer eligible women, they were all contacted for recruitment. If there were more pregnant women than the minimum required number of pregnant women, we randomly selected the required number. Then, 470 sample of pregnant women living with vulnerabilities were considered for final baseline data. This study analyzed two data sets: data prepared for sampling frame (n=1189) and data collected for baseline (n=470). The exploratory and confirmatory factor analysis used data from sampling frame, whereas associations of vulnerability to outcome measures (ANC and health facility delivery) were carried out using the baseline data.

### Item Development Procedures

Adapting Project Pathways’ population-based vulnerability segmentation approach ^31^, we conducted an extensive desk review and secondary data analysis using the 2019 Ethiopia Demographic Health Survey. The data analysis was undertaken to understand context specific drivers of maternal vulnerability affecting uptakes of health services for pregnant women. This secondary analysis identified the predictors of maternal health service utilization. These included women’s illiteracy, distance from health facilities, high parity, decision making power, exposure to media, and lack of household assets. A multivariate cluster analysis (K-medoids) was performed to assess these predictors’ ability to segment households and to classify individual pregnant women as low, moderate, and highly vulnerable. The K-medoids clustering algorithm can measure distance in multiple dimensions, representing a number of different categories or variables ^32^.

We then developed a 20-item structured questionnaire from the predictors to serve as a maternal vulnerability segmentation tool (MVST). The MVST encompasses three dimensions: economic drivers (8 items), social drivers (7 items), and drivers of information and service access (5 items). The questionnaire included individual, family, and service-related or environmental determinants. Individual-level factors included women’s literacy, work status, and ownership of a mobile phone. Family-level factors included household income, ownership of assets such as livestock and land, ownership of radio, number of children, husband’s educational status, household experience of food shortage, household decision-making autonomy, and husband’s engagement in household chores. Service-related factors included visits from health extension workers (HEWs), distance from heath facility, and residence in pre-urban or rural areas. The tool was pretested to confirm its feasibility. The 20-items questionnaire applied to our context is shown in Table1.

### Item Reduction Process

We applied exploratory factor analysis (EFA) to create reliable latent variables (factors). Orthogonal varimax type rotation with level of factor loading (> 0.3) was used to interpret factors that depicted correlation between observed variables and the factor ^33^. In the EFA, the eigenvalues and scree plot were considered to select the optimal number of factors. Correlation coefficients between variables were used to reveal the magnitude and direction of relationships. Both factor analysis and principal component analysis (PCA) are item reduction techniques. Our decision to use either of the two is based on the unique values. We found the principal component method inappropriate because the unique values were not zero for each item ^34^.

For this analysis, chi-square values cannot be considered as a reliable measure of fit with the large sample size of 1189, since χ^2^ could be inflated by large sample size ^35^. Considering our binary nature of data along with large samples and items, root mean square error of approximation (RMSEA) values of 0.01 to 0.08 ^36,37^, comparative fit index (CFI) greater than 0.90 ^35^, and Tucker Lewis index (TLI) value >0.9 ^35^ criterion were adopted to assess acceptable structural equation model and goodness-of-fit. For sampling adequacy of the EFA, we used the Kaiser–Meyer– Olkin (KMO) test (KMO>0.50) and Bartlett’s test of sphericity with p value *<*0.05 ^38^.

### Internal Reliability Test and Threshold

Different methods and cut-off points for estimation of internal reliability coefficient exist in literature. A generally accepted rule is that Cronbach alpha coefficient of 0.60-0.95 is designated as an acceptable level of reliability with a corrected item-total correlation greater than 0.3 ^39–41^. Moreover, the Kuder Richardson (known as KR-20 formula), an equivalent of Cronbach alpha statistic for binary items, could be an alternative method to assess internal consistency of the items with a level of > 0.5 suggesting good reliability coefficient ^42^.

Evidence suggests that a scale in the preliminary stages of development is generally not thought to require the reliability of one used to discriminate between groups or of one being used to make decisions about individual segmentation ^43^. A study suggested that researchers must be mindful of context, as acceptable values of reliability depend on the purpose for which the test is being used ^37^. Considering these suggestions along with our binary data, we used both Cronbach alpha and the Kuder-Richardson KR-20 coefficient criteria: having a level of 0.60 or greater internal reliability with a corrected item-total correlation greater than 0.3. Items with internal reliability were explored for pre-urban, rural and urban-rural settings.

Finally, an odds of trend analysis was conducted to test the relationship between vulnerability scores and health facility delivery. Items with acceptable internal reliability for urban-rural and rural settings were tested to confirm association with antenatal care and institutional delivery through mediation regression analysis to assess the direct and indirect effects of vulnerability scores and antenatal care on health facility delivery. In the mediation regression analysis, we controlled other potential confounders (Figure 4). However, we didn’t do prediction analysis for pre-urban setting because of small cases. All analyses were performed using STATA version 18 software. Our goal was to create and test the maternal vulnerability segmentation tools to determine the extent to which can serve as a practical and useful audience segmentation device.

## Results

### Characteristics of Respondents

A total of 1,189 households were assessed using our maternal vulnerability segmentation tool. The assessment was conducted through interviews with pregnant women in each household. Table 1 shows the proportion of households across these maternal vulnerability segmentation items. Most of the interviewed pregnant women (83.1%) lived in rural areas. Almost all households (98.3%) were male headed. Majority of the interviewed women (75.4%) could not read or write and did not work for income. A majority (77.0%) of the women had no functional mobile phone.

**Table 1.** Percent of responses by study participants to each item (n=1,189)

| Item code | Questions/items | Response options | Percent |
| --- | --- | --- | --- |
| vsq1 | Place of residence | Semi-urban | 16.9 |
|  |  | Rural | 83.1 |
| vsq2 | Head of the household | My husband (men) | 98.3 |
|  |  | Myself (women) | 1.7 |
| vsq3 | Marital status | Married | 98.3 |
|  |  | Single/dissolved | 1.7 |
| vsq4 | Women occupation | Working for income | 24.6 |
|  |  | Not working for income | 75.4 |
| vsq5 | Husband occupation | Working for income | 92.8 |
|  |  | Not working for income | 7.2 |
| vsq6 | Women education | Read and write | 24.6 |
|  |  | Not read and write | 75.4 |
| vsq7 | Husband education | Read and write | 62.8 |
|  |  | Not read and write | 37.2 |
| vsq8 | Number of children | Less than 4 | 75.5 |
|  |  | Greater or equal to 4 | 24.5 |
| vsq9 | Size of land for farming | More than 2 *Timad* | 35.3 |
|  |  | Two or less *Timad* | 64.7 |
| vsq10 | House made from | Corrugated iron | 45.3 |
|  |  | Not corrugated iron | 54.7 |
| vsq11 | Livestock ownership | Have livestock | 37.8 |
|  |  | Have no livestock | 62.2 |
| vsq12 | Household mobile phone ownership | Having mobile at home | 81.3 |
|  |  | Have no mobile at home | 18.7 |
| vsq13 | Women mobile phone ownership | Women have mobile | 23.0 |
|  |  | Women have no mobile | 77.0 |
| vsq14 | Household radio ownership | Have radio | 13.9 |
|  |  | Have no radio | 86.1 |
| vsq15 | Money to buy medical drugs | Have money | 47.6 |
|  |  | Have no money | 52.4 |
| vsq16 | Household face shortage of food in the past 3 months | No shortage of food | 64.5 |
|  |  | Experienced shortage | 35.5 |
| vsq17 | Home visits by health extension workers in the past six months | Visited by HEWs | 12.0 |
|  |  | Not visited by HEWs | 88.0 |
| vsq18 | Distance to the health facility | Near to health facility <30m | 49.6 |
| | | Far from health facility $\geq 30m$ | 50.4 |
| vsq19 | Decision-making to purchase items | Decide myself | 52.9 |
|  |  | Decide by husband | 47.1 |
| vsq20 | Husband support of household chores | Have husband support | 38.0 |
|  |  | No husband support | 62.0 |

Note: All answer choices signifying greater wealth and facilitating favors were coded as 0 and factors contributing to vulnerability were coded as 1.

More than half of the households (62.2%) had no livestock. Nearly a third of the households (35.5%) experienced food shortages over the past three months. Most households (88.0%) have not received a visit from a health extension worker in the past six months. Almost half of the women (50.4%) lived more than 30 minutes away from a health facility. Additionally, in nearly half of the households (47.1%), only the husband decided on the purchase of major household items. About one in five households (18.7%) had a mobile phone owned by any household member. Approximately a third of husbands (37.2%) could not read or write. A majority of women (62.0%) reported that their husbands did not assist them with household chores.

The outputs from factor analysis using scree plot indicated that only one factor had a high eigenvalue of one. We retained four constructs for urban-rural setting and rural residents with theoretical and practical relevance and good-to-strong reliability for the vulnerability segmentation tool. As shown in Table 2, four items had factor-loadings above 0.3. These items identified for urban-rural settings include women’s rural residence, houses not made of corrugated iron, women’s didn’t have a mobile phone, and far distance from the health facility. These items provided an average inter-item correlation of 0.26 and internal reliability coefficient (Cronbach’s alpha) of 0.60. Further, we analyzed for rural setting and revealed six items with a factor loading above 0.3 including: women and husband education, houses not made of corrugated iron, women’s mobile phone ownership, household’s mobile phone ownership, and far distance from the health facility. These items provided an average inter-item correlation of 0.20, while the internal reliability coefficient (alpha) reached 0.60. In our analysis of a structural equation model (SEM) with six items for rural, four items for pre-urban, and four in urban-rural settings (Figure 1), the goodness of fit statistics was within an acceptable range: Root Mean Square Error of Approximation (RMSEA)=0.047, Probability RESMEA=0.017, comparative fit index (CFI)=0.99, and Tucker-Lewis Index (TLI)= 0.97 for all items in pre-urban and urban-rural settings and were significantly correlated at *p*<0.001. Similarly, the goodness of fit statistics for six items were found within the acceptable range: RMSEA=0.057, CFI=0.94, and TLI=0.90. Thus, the confirmatory test findings suggest that the four and six items provided an acceptable level of factor creation in all tests.

**Figure 1:**
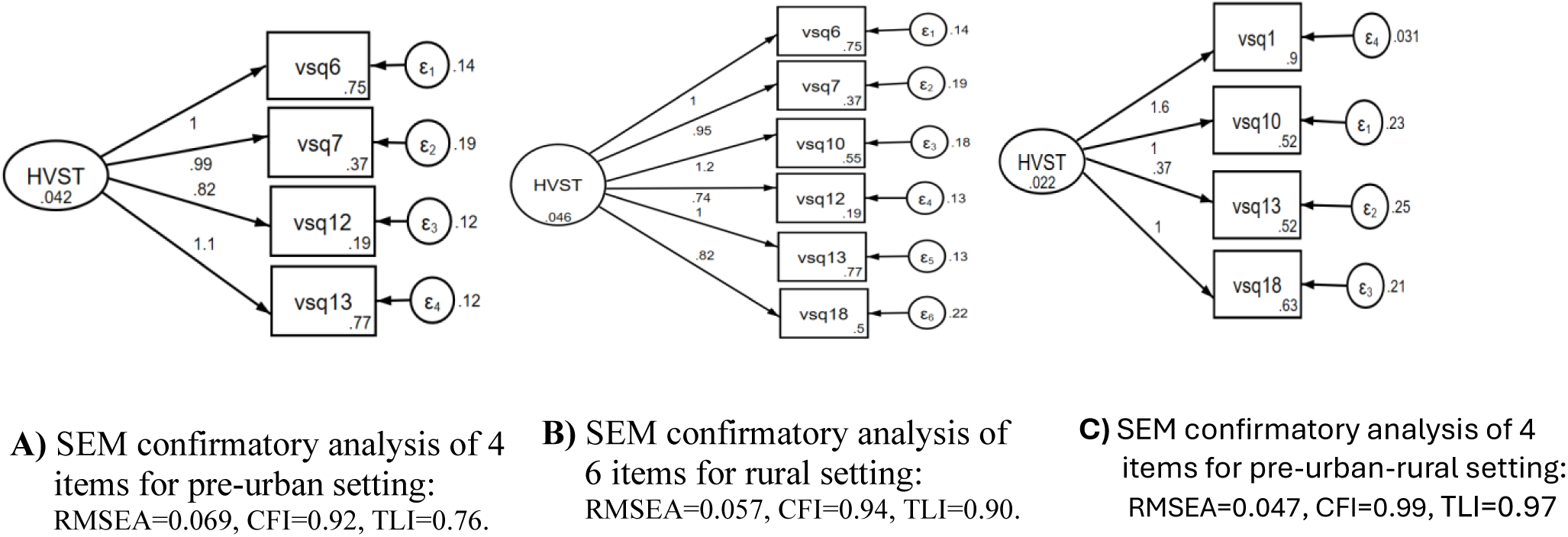
Confirmatory factor analysis using structural equation modeling (SEM) items retained after rotating 4 items for pre-urban indicated in A, and 6 items for rural setting indicated in B and 4 items for urban-rural setting indicated in C (n=1,189).

**Table 2:** Reduced items with internal reliability coefficients using factor analysis for rural, semi urban and urban-rural settings (n=1,189)

| Retained items after rotation | Item loading | Internal reliability statistics |  |  |  |  |
| --- | --- | --- | --- | --- | --- | --- |
|  | Factor Loadings >0.3 | Inter item correlation | Item-rest correlation | Average interitem correlation | Cronbach alpha coefficient | KR20 coefficient |
| **Rural setting** |  |  |  |  |  |  |
| Women education | 0.47 | 0.60 | 0.37 | 0.20 | 0.55 |  |
| Husband education | 0.42 | 0.57 | 0.33 | 0.21 | 0.56 |  |
| House made from | 0.50 | 0.61 | 0.38 | 0.19 | 0.54 |  |
| Women mobile ownership | 0.41 | 0.56 | 0.31 | 0.21 | 0.57 |  |
| Mobile ownership in home | 0.52 | 0.61 | 0.38 | 0.19 | 0.54 |  |
| Distance to health facility | 0.33 | 0.52 | 0.27 | 0.22 | 0.59 |  |
| **Test scale** |  |  |  | **0.20** | **0.61** | **0.60** |
| **Semi-urban setting\*** |  |  |  |  |  |  |
| Women education | 0.45 | 0.62 | 0.29 | 0.23 | 0.48 |  |
| Husband education | 0.43 | 0.66 | 0.34 | 0.20 | 0.43 |  |
| Mobile ownership in home | 0.35 | 0.58 | 0.22 | 0.27 | 0.53 |  |
| Women mobile ownership | 0.60 | 0.71 | 0.42 | 0.16 | 0.36 |  |
| **Test scale** |  |  |  | **0.22** | **0.52** | **0.53** |
| **Urban-Rural setting** |  |  |  |  |  |  |
| Place of residence | 0.60 | 0.73 | 0.46 | 0.21 | 0.44 |  |
| House made from | 0.65 | 0.72 | 0.45 | 0.22 | 0.45 |  |
| Women mobile ownership | 0.37 | 0.62 | 0.30 | 0.31 | 0.57 |  |
| Distance to health facility | 0.35 | 0.61 | 0.28 | 0.32 | 0.58 |  |
| **Test scale** |  |  |  | **0.26** | **0.60** | **0.60** |

\*We refer to semi-urban means a district town resident and because of small semi-urban cases we did not do prediction analysis

Figure 2 shows the percentage of health facility delivery and odds of trend with the composite scores of four and six items in urban-rural and rural settings. In Figure 3A and 3B, women in low vulnerability groups had higher percentage of health facility delivery compared to those in higher vulnerability groups. As shown in Figure 2C and 2D, we observed a declining trend in health facility delivery with an increase in vulnerability composite score of four and six item points starting from intersecting score of 3 points (*p*<0.001). These findings indicate that the four and six items can segment pregnant women in urban-rural and rural settings, respectively, for women who are at higher risk of institutional delivery. For health facility delivery as the outcome of interest, a cut-off points of less than 3 (or below) showed greater odds of institutional delivery, whereas those scoring above 3 were more likely to deliver at home.

**Figure 2:**
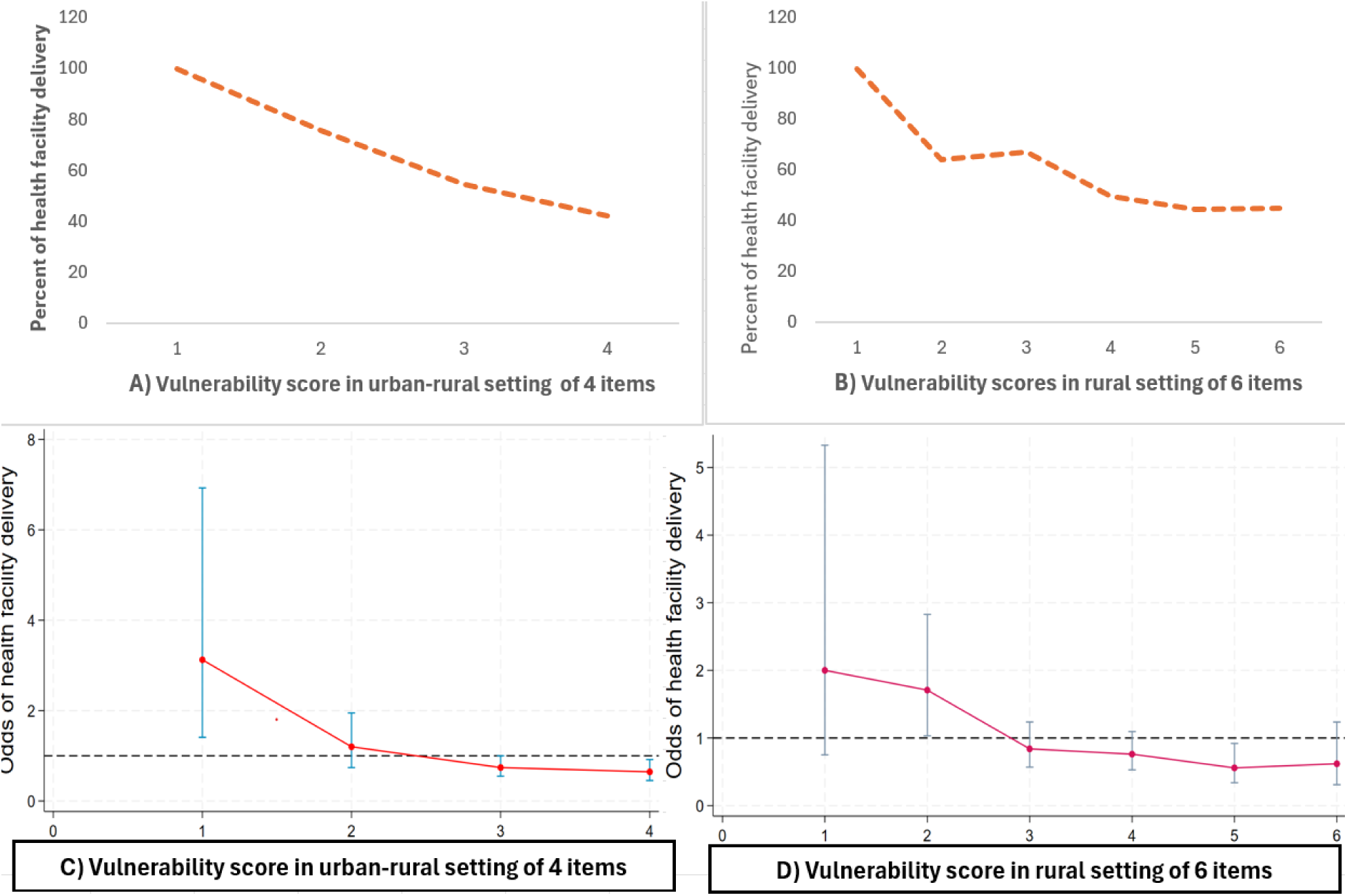
Odds of trend relationship between ***health facility delivery*** and vulnerability score from urban-rural women indicated in figure A and C and rural women indicated in figure B and D.

**Figure 3:**
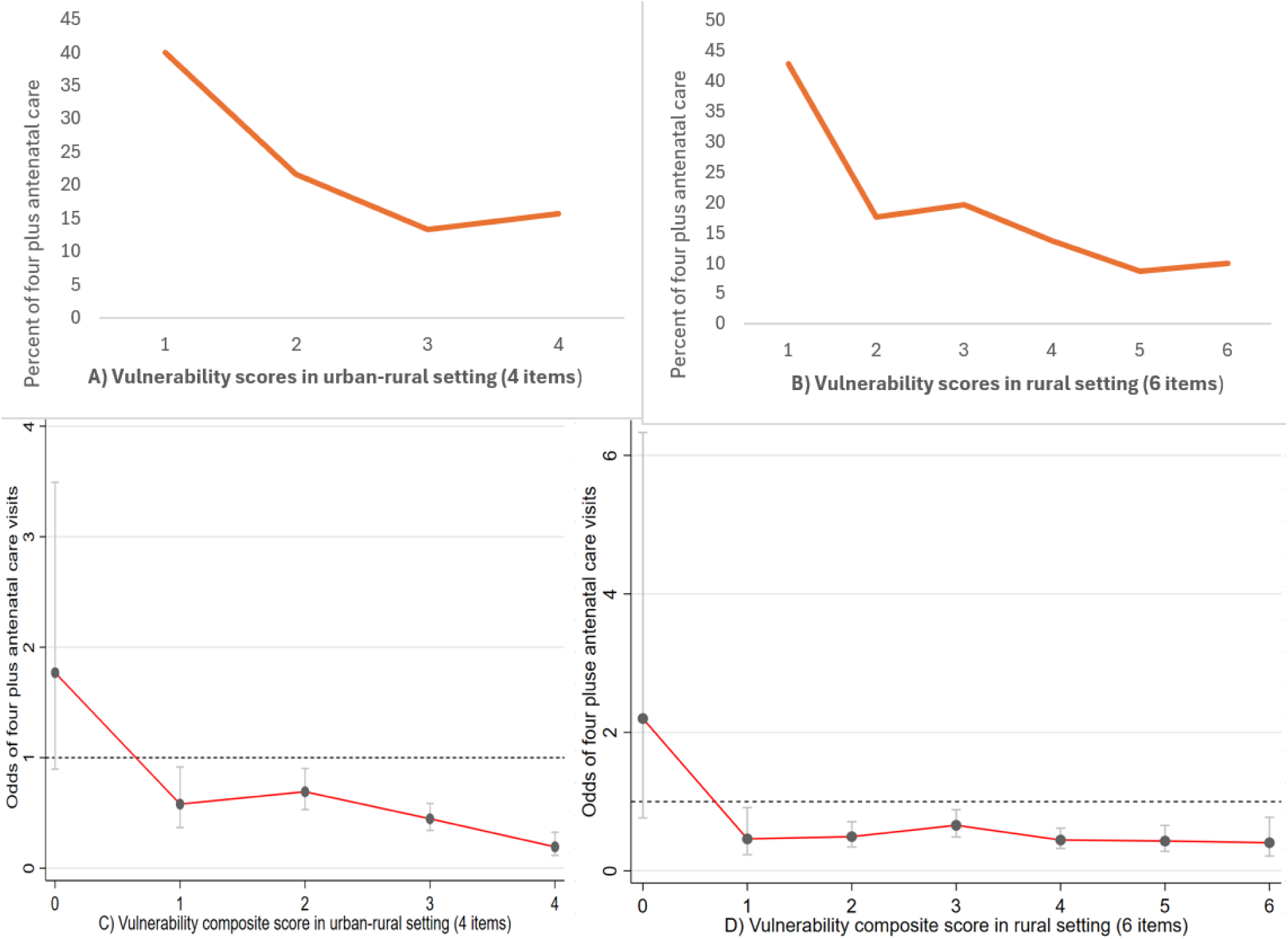
Odds of trend relationship between ***antenatal care (ANC)*** and vulnerability score from urban-rural women indicated in figure A and C and rural women indicated in figure B and D.

Similarly, Figure 3 shows percentage of antenatal care utilization and odds of trend with composite scores of four and six items compared with urban-rural and rural settings. In Figure 3A and 3B, women in low vulnerability groups had higher percentage of antenatal care visits, compared to higher vulnerability groups. In Figure 3C and 3D, the composite score of the four items in urban-rural setting and six items in rural setting showed a steady decline in antenatal care visits with statistically significant values among women who have greater than one or more intersecting vulnerability score points (*p*<0.001). These findings indicate that the four and six items in urban-rural and rural settings, respectively, can segment women who are attending antenatal care less than the recommended number of visits. Odds of antenatal care visits with vulnerability score showed that the cutoff point less than one indicated less vulnerability and a score of one or more indicated greater vulnerability.

A mediation regression analysis was done to depict the direct, indirect, and total effects of vulnerability indexes on maternal services. As shown in Figure 4, the effect of vulnerability on health facility delivery was mediated through antenatal care visits. This mediation could validate the odds of trend analysis shown above in Figure 2 and Figure 3.

**Figure 4:**
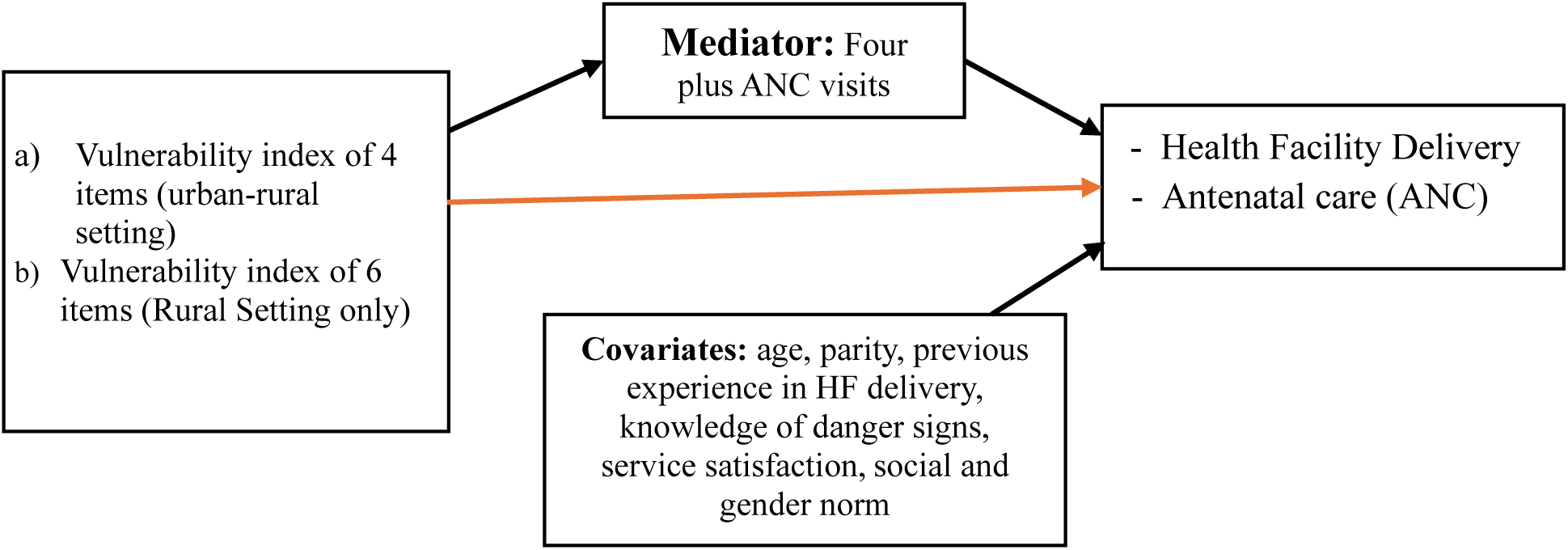
Mediators of vulnerability indexes and maternal health services with covariates to show direct, indirect and total effects.

As indicated in Figure 5A and 5C (4 items identified for urban-rural settings), women in higher vulnerability groups were less likely to go for antenatal care visits and were less likely to deliver in health facilities, as compared to women in the low vulnerability groups (*p*<0.001 of net direct effect (NDE) and total effect (TE). However, the net indirect effect (NIE) of vulnerability on health facility delivery mediating through effects of antenatal care was not significant (Figure 5 of A).

**Figure 5:**
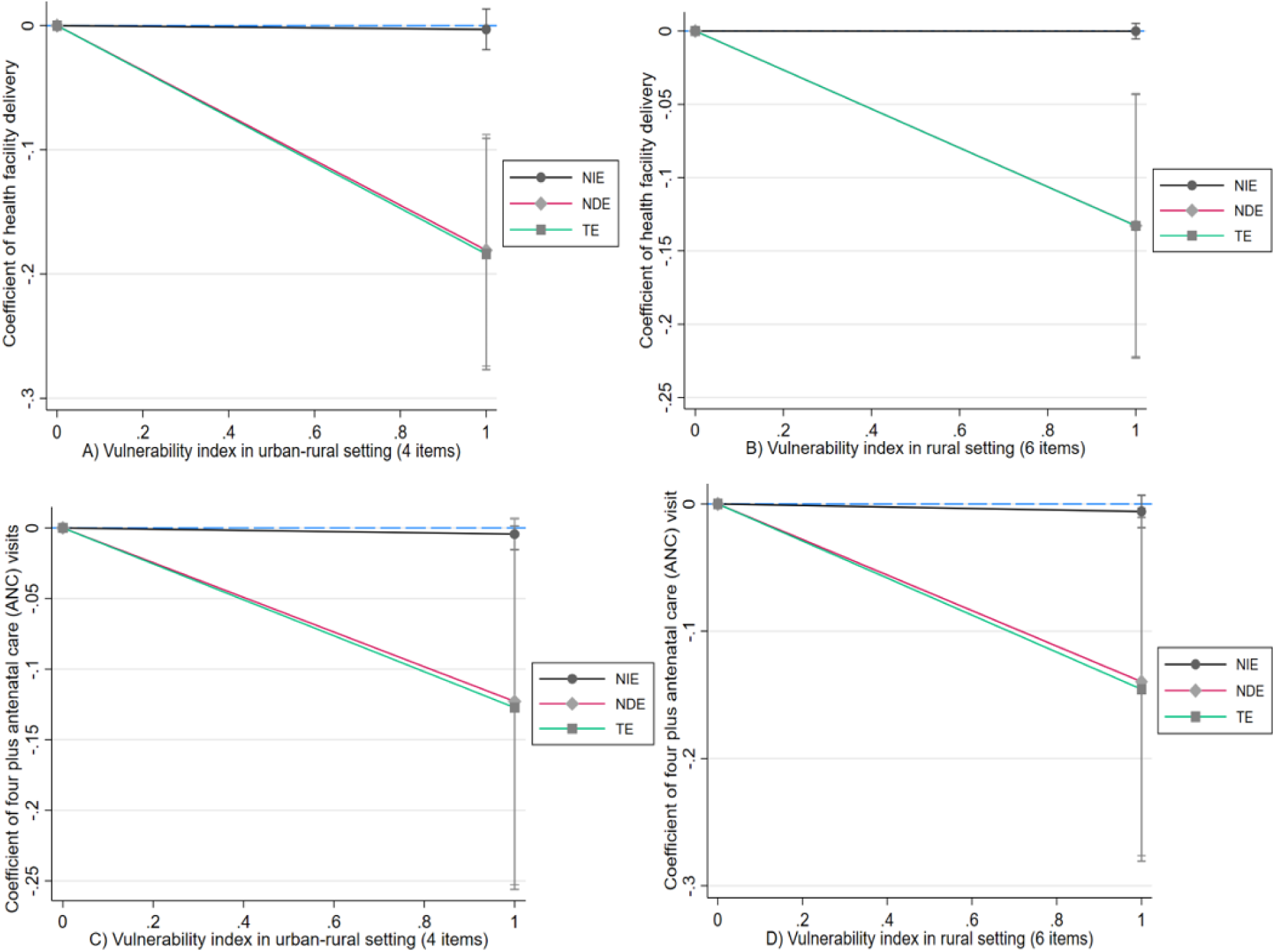
Mediation analysis between vulnerability scores and health facility delivery, and antenatal care among urban-rural and rural women indicated in A and C, and in B and D respectively.

Similarly, in Figure 5B and 5D (6 items identified for rural setting), women living with vulnerability were less likely to go for antenatal care visits and less likely to deliver in health facilities, compared to women with low vulnerabilities (*p*<0.01 of NDE and TE).

## Discussion

The maternal vulnerability segmentation tool (MVST) we introduced and tested in this paper, we believe, is both practical and valuable for a number of reasons. First, literature indicates that interventions that are more narrowly targeted are likely to be more effective and cost-effective than those that can only make a generic appeal ^44–46^. A tool like the MVST that segments audiences according to their vulnerability is a significant first step in developing tailored interventions: knowing the audience segment any given pregnant woman falls under can help interventions develop targeted materials and methods to appeal to her specifically.

Second, conceptually (though not yet practically), this tool may be able to reduce disparities in maternal mortality. In Ethiopia, like in many other low- and middle-income countries, we have seen significant progress in reducing maternal mortality ^3,7^, but even the prevailing rates are much higher than is the case in their developed-country counterparts ^1,2,47^. This is because of what we might call the *neglect at the margins* – while interventions, educational programs, and government policies have reduced the overall mortality, women at the edges of society, those who are subjected to many intersecting vulnerability factors, have not been well served by existing policies and interventions. This is likely because of the unique constellation of vulnerability-enhancing factors that they face. Evidence shows that women experience various risks and barriers during pregnancy, resulting in adverse maternal and neonatal outcomes, such as pre-eclampsia, preterm birth, and stillbirth ^48^. The MVST is designed to identify women at the highest levels of risk. While the tool, of course, does not develop specific interventions for them, it provides the first step in doing so – by identifying who the women are in a community.

Third, the VMST is meant to be an easy-to-use device that does not come with a large literacy burden. We envision that this tool will be used by frontline workers (in Ethiopia, they are often not highly educated) with relative ease to identify women who fall in the high-vulnerability category. Once they can be so identified, frontline workers can then be trained in addressing the unique challenges that the women face. Furthermore, this tool can be used in both an urban and a rural setting, places where some of the underlying challenges, vulnerability sources, and deficits can be different.

We should note, however, that the MVST is for identifying women living with vulnerabilities; it is not an intervention strategy. Put another way, it is a population segmentation tool, not a tailoring tool. Hence, the next step for us is to test the efficacy of this tool in a real world setting to learn whether it can effectively identify different population segments, and once so segmented, whether interventions can be designed to cater specifically to the different audience segments.

We acknowledge certain limitations of our study. Given our focus on identifying those most disadvantaged in accessing care, our vulnerability criteria may not be universally applicable across broader regions of Ethiopia, where societal, cultural, and environmental vulnerabilities differ from our study sites. Nevertheless, we believe the process of developing a tool to identify the most invisible populations and refining it based on residential areas to enhance its practical utility can serve as a replicable approach in other contexts. Further, we acknowledge that maternal vulnerability is not a static concept. Rather, women’s levels and facets of vulnerability continuously evolve throughout the stages of pregnancy, childbirth, and the postpartum period, all of which are influenced by pre-existing health conditions and their responses to emerging threats and barriers. Future research could benefit from refining our tool to capture vulnerability-enhancing factors unique to the different stages of pregnancy. Lastly, our study focused on maternal care-seeking behaviors as outcomes, emphasizing its potential to address gaps in service coverage. While we consider service utilization a strong predictor of maternal and child outcomes, we recommend testing the MVST against health outcomes to uncover additional important vulnerability factors.

To our knowledge, this is the first study that consolidates holistic factors affecting maternal health service utilization to develop a tool that identifies the most underserved pregnant women in Ethiopia. While other vulnerability tools are extant, we believe that the MVST offers a targeted perspective by taking account of the health system’s contexts along with the sociocultural dimensions relevant to our target population. The MVST also ensures its practicality in the field, as it presents only the essential factors that define women’s vulnerability and fine tunes its application based on residential areas. This approach results in a tool that can be used to inform effective, cost-saving intervention strategies, while minimizing the burden on frontline workers and pregnant women during roll-out. It would be worth further developing and validating vulnerability segmentation tools for maternal and child health programing.

## Data Availability

Not yet

## Acknowledgements

We would like to acknowledge all pregnant women and women development armies for their active participation during recruitment process. The authors acknowledge CCP staffs who involved in the recruitment and implementation process including Tewabech Tesfalegn, Minyahil Ayele, Melese Jenbolla, Dr. Jordan Asheafi, Dr.Tsega Endrias, Shewaye Tsegaye, and all others who played a major role. We also thank Paula Stauffer and Sanjanthi Velu for their overall supervision and technical support throughout the project phase. Finally, we thank the Bill and Melinda Gates Foundation for providing the financial and technical support needed.

## Declarations Ethics approval

The study received approval by the Ethiopian Public Health Institute (EPHI), a governmental public health institution located in Addis Ababa, Ethiopia (EPHI-IRB-510-2023). The study was also approved by the (*** University; masked for blind review) Institutional Review Board. For vulnerability segmentation practice, we obtained another ethical approval for public health practice by the (XXX University) Institutional Review Board (IRB00023366).

## Authors’ contributions

Lakew Y led the conceptualization, study design, analysis and writing of the manuscript. Habtamu Tamene and Rajiv Rime contributed to the conceptualization, validation of data analysis and writing part of the manuscript. Kang BA, Kapadia N and Kuka S.H are involved in the study design and critical review.

## References

1. Mgawadere F, Kana T, van den Broek N. Measuring maternal mortality: a systematic review of methods used to obtain estimates of the maternal mortality ratio (MMR) in low- and middle-income countries. Br Med Bull. 2017 Jan 1;121(1):121–34.

2. WHO. TRENDS IN MATERNAL MORTALITY: 2000 TO 2017, Estimates by WHO, UNICEF, UNFPA, World Bank Group and the United Nations Population Divisie [Internet]. Licence: CC BY-NC-SA 3.0 IGO; 2019. Available from: https://www.unfpa.org/sites/default/files/pub-pdf/Maternal_mortality_report.pdf

3. WHO. Trends in maternal mortality 2000 to 2020: estimates by WHO, UNICEF, UNFPA, World Bank Group and UNDESA/Population Division [Internet]. 2023. Available from: https://www.who.int/publications/i/item/9789240068759

4. Sule FA, Uthman OA, Olamijuwon EO, Ichegbo NK, Mgbachi IC, Okusanya B, et al. Examining vulnerability and resilience in maternal, newborn and child health through a gender lens in low-income and middle-income countries: a scoping review. BMJ Glob Health. 2022 Apr;7(4):e007426.

5. Roro EM, Tumtu MI, Gebre DS. Predictors, causes, and trends of neonatal mortality at Nekemte Referral Hospital, east Wollega Zone, western Ethiopia (2010–2014). Retrospective cohort study. Ameh CA, editor. PLoS ONE. 2019 Oct 9;14(10):e0221513.

6. Hasan MM, Magalhaes RJS, Fatima Y, Ahmed S, Mamun AA. Levels, Trends, and Inequalities in Using Institutional Delivery Services in Low- and Middle-Income Countries: A Stratified Analysis by Facility Type. Glob Health Sci Pract. 2021 Mar 31;9(1):78–88.

7. Central Statistical Agency - CSA/Ethiopia, ICF. Ethiopia Demographic and Health Survey 2016. 2017. [Internet]. Available from: https://dhsprogram.com/pubs/pdf/fr328/fr328.pdf

8. Central Statistical Agency - CSA/Ethiopia, ICF. Ethiopia Demographic and Health Survey 2019 [Internet]. Available from: https://dhsprogram.com/pubs/pdf/FR363/FR363.pdf

9. Banteyerga H. Ethiopia’s health extension program: improving health through community involvement. MEDICC Rev. 2011 Jul;13(3):46–9.

10. WHO. WHO Recommendations on Antenatal Care for a Positive Pregnancy Experience: Summary 2016. [Internet]. Available from: https://apps.who.int/iris/bitstream/handle/10665/259947/WHO-RHR-18.02-eng.pdf

11. Carlo WA, Goudar SS, Jehan I, Chomba E, Tshefu A, Garces A, et al. Newborn-care training and perinatal mortality in developing countries. N Engl J Med. 2010 Feb 18;362(7):614–23.

12. Randive B, San Sebastian M, De Costa A, Lindholm L. Inequalities in institutional delivery uptake and maternal mortality reduction in the context of cash incentive program, Janani Suraksha Yojana: results from nine states in India. Soc Sci Med. 2014 Dec;123:1–6.

13. Wong KLM, Restrepo-Méndez MC, Barros AJD, Victora CG. Socioeconomic inequalities in skilled birth attendance and child stunting in selected low and middle income countries: Wealth quintiles or deciles? PLoS One. 2017;12(5):e0174823.

14. Darmstadt GL, Bhutta ZA, Cousens S, Adam T, Walker N, de Bernis L, et al. Evidence-based, cost-effective interventions: how many newborn babies can we save? Lancet. 2005 Mar 12;365(9463):977–88.

15. Gurmesa Tura, Mesganaw Fantahun and Alemayehu Worku, Tura G etal. The effect of health facility delivery on neonatal mortality: systematic review and meta-analysis. BMC Pregnancy and Childbirth volume [Internet]. 2013; Available from: http://www.biomedcentral.com/1471-2393/13/18

16. Arora S, Shah D, Chaturvedi S, Gupta P. Defining and measuring vulnerability in young people. Indian J Community Med. 2015;40(3):193.

17. Briscoe L, Lavender T, McGowan L. A concept analysis of women’s vulnerability during pregnancy, birth and the postnatal period. J Adv Nurs. 2016 Oct;72(10):2330–45.

18. Adger WN. Social and ecological resilience: are they related? 2006. Progress in Human Geography. 2000 Sep;24(3):347–64.

19. Vulnerability: A View from Different Disciplines. Jeffrey Alwang, Paul B. 2001 [Internet]. Available from: https://documents1.worldbank.org/curated/en/636921468765021121/pdf/multi0page.pdf

20. Adger WN. Vulnerability.Global Environmental Change 2001. Global Environmental Change. 2006 Aug;16(3):268–81.

21. Grigorescu I, Mocanu I, Mitrică B, Dumitraşcu M, Dumitrică C, Dragotă CS. Socio-economic and environmental vulnerability to heat-related phenomena in Bucharest metropolitan area. Environ Res. 2021 Jan;192:110268.

22. Adhav CA, R S, Chandel BS, Bhandari G, Ponnusamy K, Ram H. Socio-economic vulnerability to climate change – Index development and mapping for districts in Maharashtra, India. SSRN Journal [Internet]. 2021 [cited 2024 Oct 17]; Available from: https://www.ssrn.com/abstract=3854297

23. Johnson DP, Stanforth A, Lulla V, Luber G. Developing an applied extreme heat vulnerability index utilizing socioeconomic and environmental data. Applied Geography. 2012 Nov;35(1–2):23–31.

24. Surgo Ventures. Building a Maternal Vulnerability Index for the United States [Internet]. Available from: https://surgoventures.org/portfolio/action-areas/building-a-maternal-vulnerability-index-for-the-united-states

25. FHI360 Vulnerability Assessment Methodologies: A Review of the Literature [Internet]. Available from: https://www.fhi360.org/wp-content/uploads/drupal/documents/Vulnerability%20Assessment%20Literature%20Review.pdf

26. Sule FA, Uthman OA, Olamijuwon EO, Ichegbo NK, Mgbachi IC, Okusanya B, et al. Examining vulnerability and resilience in maternal, newborn and child health through a gender lens in low-income and middle-income countries: a scoping review. BMJ Glob Health. 2022 Apr;7(4):e007426.

27. Makinde OA, Uthman OA, Mgbachi IC, Ichegbo NK, Sule FA, Olamijuwon EO, et al. Vulnerability in maternal, new-born, and child health in low- and middle-income countries: Findings from a scoping review. Ranabhat CL, editor. PLoS ONE. 2022 Nov 11;17(11):e0276747.

28. Theron L. Resilience of sub-Saharan children and adolescents: A scoping review. Transcult Psychiatry. 2023 Dec;60(6):1017–39.

29. Naudé W, Santos-Paulino AU, McGillivray M. Measuring Vulnerability: An Overview and Introduction. Oxford Development Studies. 2009 Sep;37(3):183–91.

30. Kang BA, Tamene H, Lakew Y, Stephens D, Rimal R. Developing and evaluating human-centered design solutions for enhancing maternal health service utilization among vulnerable pregnant women in Oromia, Ethiopia: Study protocol for a quasi-experimental study. Gates Open Res. 2024 Dec 18;8:93.

31. Johnson TP, Wendland M. A Role for Design in Global Health: Making the Concept of Vulnerability Actionable. She Ji: The Journal of Design, Economics, and Innovation. 2022;8(4):486–503.

32. Qualtrics XM for Strategic Research. [Internet]. What is cluster analysis? Overview and examples. Available from: https://www.qualtrics.com/experience-management/research/cluster-analysis/

33. Hair J.F., Jr., Black W.C., Babin B.J., Anderson R.E. Multivariate Data Analysis. 7th ed. Prentice Hall; Upper Saddle River, NJ, USA: 2010. In. Available from: https://www.drnishikantjha.com/papersCollection/Multivariate%20Data%20Analysis.pdf

34. Acock, Alan C. (2016): A Gentle Introduction to Stata. Fifth Edition. TX: Stata Press. In. Available from: https://www.stata-press.com/books/preview/acock6-preview.pdf

35. Brown, T. A. (2015). Confirmatory factor analysis for applied research. 2nd edition. Guilford publications. [Internet]. Available from: file:///C:/Users/yihuniel/Downloads/MethodologyintheSocialSciencesTimothyA.BrownPsyD-ConfirmatoryFactorAnalysisforAppliedResearchSecondEdition-TheGuilfordPress2015%20(1).pdf

36. Byrne BM. Structural equation modeling with Mplus: Basic concepts, applications, and programming. New York: Routledge, 2012. Available from: https://www.researchgate.net/publication/319269176_Structural_equation_modeling_with_Mplus_Basic_concepts_applications_and_programming_New_York_Taylor_FrancisRoutledge

37. Muthen LK, Muthen BO. Mplus user’s guide: The comprehensive modeling program for applied researchers, 2012. [Internet]. Available from: https://socialwork.wayne.edu/research/pdf/mplus-users-guide.pdf

38. Jacobs NW, Berduszek RJ, Dijkstra PU, van der Sluis CK. Validity and Reliability of the Upper Extremity Work Demands Scale. J Occup Rehabil. 2017 Dec;27(4):520–9.

39. Ursachi G, Horodnic IA, Zait A. How Reliable are Measurement Scales? External Factors with Indirect Influence on Reliability Estimators. Procedia Economics and Finance. 2015;20:679–86.

40. Taber KS. The Use of Cronbach’s Alpha When Developing and Reporting Research Instruments in Science Education. Res Sci Educ. 2018 Dec;48(6):1273–96.

41. Janssens W, editor. Marketing research with SPSS [Internet]. Nachdr. Harlow Munich: Financial Times Prentice Hall; 2011. 441 p. Available from: https://api.pageplace.de/preview/DT0400.9780273732280_A24658680/preview-9780273732280_A24658680.pdf

42. Blunch, N. J. (2008). Introduction to structural equation modelling using SPSS and AMOS. Sage Publications Ltd [Internet]. Available from: https://methods.sagepub.com/book/intro-to-structural-equation-modelling-using-spss-amos

43. Robert A. Peterson. A Meta-Analysis of Cronbach’s Coefficient Alpha. Journal of Consumer Research Vol. 21, No. 2 (Sep., 1994), pp. 381–391 (11 pages) Published By: Oxford University Press. Available from: https://www.jstor.org/stable/2489828

44. Ryan P, Lauver DR. The Efficacy of Tailored Interventions. J of Nursing Scholarship. 2002 Dec;34(4):331–7.

45. Noar SM, Benac CN, Harris MS. Does tailoring matter? Meta-analytic review of tailored print health behavior change interventions. Psychological Bulletin. 2007;133(4):673–93.

46. Ishikawa Y, Hirai K, Saito H, Fukuyoshi J, Yonekura A, Harada K, et al. Cost-effectiveness of a tailored intervention designed to increase breast cancer screening among a non-adherent population: a randomized controlled trial. BMC Public Health. 2012 Dec;12(1):760.

47. Global, regional, and national levels of maternal mortality,1990–2015: a systematic analysis for the Global Burden of Disease Study 2015. Lancet 2016; 388: 1775–812 [Internet]. Available from: https://www.thelancet.com/action/showPdf?pii=S0140-6736%2816%2931470-2

48. Sheikh J, Allotey J, Kew T, Khalil H, Galadanci H, Hofmeyr GJ, et al. Vulnerabilities and reparative strategies during pregnancy, childbirth, and the postpartum period: moving from rhetoric to action. eClinicalMedicine. 2024 Jan;67:102264.

